# The Association Between Obstructive Sleep Apnea Defined by 3 Percent Oxygen Desaturation or Arousal Definition and Self-Reported Cardiovascular Disease in the Sleep Heart Health Study

**DOI:** 10.1101/2020.09.22.20199745

**Authors:** Stuart F. Quan, Rohit Budhiraja, Sogol Javaheri, Sairam Parthasarathy, Richard B. Berry

## Abstract

**Background:** Studies have established that OSA defined using a hypopnea definition requiring a ≥4% oxygen desaturation (AHI4%) is associated with cardiovascular (CVD) or coronary heart (CHD) disease. This study determined whether OSA defined using a hypopnea definition characterized by a ≥3% oxygen desaturation or an arousal (AHI3%A) is associated with CVD/CHD.

**Methods:** Data were analyzed from 6307 Sleep Heart Health Study participants who had polysomnography. Self-reported CVD included angina, heart attack, heart failure, stroke or previous coronary bypass surgery or angioplasty. Self-reported CHD included the aforementioned conditions but not stroke or heart failure. The association between OSA and CVD/CHD was examined using logistic regression models with stepwise inclusion of demographic, anthropometric, social/behavioral and co-morbid medical conditions. A parsimonious model in which diabetes and hypertension were excluded because of their potential to be on the causal pathway between OSA and CVD/CHD also was constructed.

**Results:** For CVD, the odds ratios and 95% confidence intervals for AHI3%A ≥30/hour were 1.39 (1.03-1.87) and 1.45 (1.09-1.94) in the fully adjusted and parsimonious models. Results for CHD were 1.29 (0.96-1.74) and 1.36 (0.99-1.85). In participants without OSA according to more stringent AHI4% criteria but with OSA using the AHI3%A definition, similar findings were observed.

**Conclusion:** OSA defined using an AHI3%A is associated with both CVD and CHD. Use of a more restrictive AHI4% definition will misidentify a large number of individuals with OSA who have CVD or CHD. These individuals may be denied access to therapy, potentially worsening their underlying CVD or CHD.

## Introduction

Obstructive sleep apnea (OSA) is a common disorder characterized by recurrent episodes of either complete upper airway collapse (apneas) or partial collapse (hypopneas) during sleep. A number of large studies have established that OSA is a risk factor for the development of hypertension and cardiovascular disease (CVD) as well as higher mortality; individuals with more severe OSA are at greater risk (1-3). The most commonly used metric of OSA severity is the apnea hypopnea index (AHI). However, there is controversy regarding the definition of the AHI. In 2012, the American Academy of Sleep Medicine (AASM) recommended that the hypopnea definition include any decrease in airflow by at least 30% from the baseline with an oxyhemoglobin desaturation of at least 3%, or an arousal from sleep (4). However, several payors including the Centers for Medicare and Medicaid Services (CMS) continue to require a more stringent hypopnea definition necessitating a 4% or greater decrease in oxygen saturation (5) despite evidence documenting a relationship between the AASM recommended standard and daytime sleepiness (6). The resistance to universal acceptance of the AASM criteria is based in part on the lack of evidence that 3% desaturations or arousals have an adverse cardiovascular impact. This reluctance to adopt a more inclusive definition of sleep apnea has restricted access to OSA treatment for many patients (7). Therefore, determining if there is relationship between OSA characterized by at least 3% drop in saturation or an arousal from sleep and CVD may assist in identification of persons at risk for CVD, allow greater access to care and potentially improve other health-related outcomes.

Using the database from the Sleep Heart Health Study, a large well-characterized community based cohort that had undergone polysomnography, the current study aimed to determine the association between the AASM recommended definition of the AHI which incorporates hypopneas with at least a 3% desaturation or an arousal (AHI3%A) and self-reported CVD and coronary heart disease (CHD) in middle-aged and older adults. In addition, we sought to ascertain whether there was an association between CVD or CHD and OSA severity among individuals who were not identified as having OSA using the more restrictive standard of requiring at least a 4% oxygen desaturation irrespective of an arousal (AHI4%), but were classified as having OSA by the AHI3%A definition. We hypothesized that increasing OSA severity represented by the AHI3%A would be associated with a greater likelihood of having prevalent CVD or CHD, and that persons who were not identified as having OSA using the AHI4% criteria would have a higher likelihood as well.

## Methods

This study analyzed data obtained from the Sleep Heart Health Study (SHHS) which was a prospective multicenter cohort study designed to investigate the relationship between OSA and cardiovascular diseases in the United States. The study’s rationale and design have been published elsewhere (8). Briefly, 6,441 subjects, 40 years of age and older were recruited starting in 1995 from several ongoing “parent” cardiovascular and respiratory disease cohorts that were initially assembled between 1976 and 1995 (9). These cohorts included the Offspring Cohort and the Omni Cohort of the Framingham Heart Study in Massachusetts; the Hagerstown, MD, and Minneapolis, MN, sites of the Atherosclerosis Risk in Communities Study; the Hagerstown, MD, Pittsburgh, PA, and Sacramento, CA, sites of the Cardiovascular Health Study; 3 hypertension cohorts (Clinic, Worksite, and Menopause) in New York City; the Tucson Epidemiologic Study of Airways Obstructive Diseases and the Health and Environment Study; and the Strong Heart Study of American Indians in Oklahoma, Arizona, North Dakota, and South Dakota. Because of sovereignty issues, 134 participants from the Arizona cohort of the Strong Heart Study withdrew consent. Analyses were performed on the remaining 6307 participants. The SHHS was approved by each site’s institutional review board for human subjects’ research, and informed written consent was obtained from all subjects at the time of their enrollment.

### Polysomnography and Home Visit

Participants underwent overnight in-home polysomnograms using the Compumedics Portable PS-2 System (Abbottsville, Victoria, Australia) administered by trained technicians (10). The home visits were performed by two-person, mixed-sex teams in visits that lasted 1.5 to 2 hours. Participants were asked to schedule the visit so that it would occur approximately two hours prior to their usual bedtime. At the time of the home visit, an inventory of each participant’s medications was made. In addition, a health interview was completed that ascertained the presence of several health conditions. Questionnaires that were completed included the SHHS Sleep Habits Questionnaire which incorporated the Epworth Sleepiness Scale (ESS) (11) and the Medical Outcomes Study SF-36 (12). Blood pressure was measured manually in triplicate in a seated position after 5 minutes of rest (13). The average of the second and third measurements was used for this analysis. Body weight was obtained using a digital scale.

The SHHS recording montage consisted of electroencephalogram (C4/A1 and C3/A2), right and left electrooculogram, a bipolar submental electromyogram, thoracic and abdominal excursions (inductive plethysmography bands), airflow (detected by a nasal-oral thermocouple [Protec, Woodinville, WA]), oximetry (finger pulse oximetry [Nonin, Minneapolis, MN]), electrocardiogram and heart rate (using a bipolar electrocardiogram lead), body position (using a mercury gauge sensor), and ambient light (on/off, by a light sensor secured to the recording garment). Sensors were placed, and equipment was calibrated during an evening home visit by a certified technician. After technicians retrieved the equipment, the data, stored in real time on PCMCIA cards, were downloaded to the computers of each respective clinical site, locally reviewed, and forwarded to a central reading center (Case Western Reserve University, Cleveland, OH). Comprehensive descriptions of polysomnography scoring and quality-assurance procedures have been previously published (14). In brief, sleep was scored according to guidelines developed by Rechtschaffen and Kales (15). Strict protocols were maintained to ensure comparability among centers and technicians. Intra-scorer and inter-scorer reliabilities were high (14).

The apnea hypopnea index (AHI) was calculated for each participant using two definitions of hypopnea, the AASM recommended definition [AHI3%A] and the AASM acceptable [CMS] definition [AHI4%]. For AHI3%A, hypopneas were identified if the amplitude of a measure of flow or volume (detected by the thermocouple or thorax or abdominal inductance band signals) was reduced discernibly (at least 30% lower than baseline breathing) for at least 10 seconds, did not meet the criteria for apnea and the event was associated with either a 3% oxygen desaturation from baseline or terminated with electroencephalographic evidence of an arousal. For AHI4%, hypopneas were identified if the aforementioned reduction in flow or volume occurred and the event was associated with a 4% oxygen desaturation from baseline. In both cases, an apnea was defined as a complete or almost complete cessation of airflow, as measured by the amplitude of the thermocouple signal, lasting at least 10 seconds.

### Outcome Assessment

Self-reported CVD and CHD were the outcomes of interest for this analysis and were obtained from the standardized health interview performed at the time of each participant’s polysomnography home visit. Participants were asked if they had ever been told by a doctor that she or he had angina, heart attack, heart failure, or stroke and if the participant had ever undergone coronary bypass surgery or coronary angioplasty. Prevalent CVD was defined as a positive response to one or more of the aforementioned conditions or procedures. Prevalent CHD was defined as an affirmative response to the same questions with the exclusion of responses to the presence of heart failure or stroke.

### Covariates

Selection of potential covariates was based on previous studies documenting an association with either CVD or CHD. These included various demographic (e.g., sex, race/ethnicity, education, marital status), anthropometric (e.g., height, weight and blood pressure [BP]), social/behavioral (e.g., smoking history, alcohol use, sleep duration, quality of life) indices as well as plasma lipids (cholesterol, high density lipoprotein [HDL], triglycerides), several diseases (depression, hypertension, diabetes) and spirometry.

The following definitions were used for those covariates that were not primarily recorded. Body mass index (BMI) was calculated as weight (kg)/height (m^2^). The ankle arm index (AAI) was computed as the ratio of blood pressure at the ankle to that in the arm. Waist to hip ratio was the waist divided by hip circumferences. Hypertension was defined as a self-report of hypertension or the use of anti-hypertensive medications. Diabetes was considered present if it was self-reported by the participant or if there was use of oral hypoglycemic agents or insulin. Depression was defined as present if the participant indicated on the SF-36 that he/she was feeling “blue” or “down” for at least “a good bit of the time” for the previous 4 weeks, or he/she was using antidepressant medications. Insomnia was defined as often or almost always having “trouble falling asleep”, “waking up during the middle of the night and having difficulty getting back to sleep” or “waking up too early in the morning and being unable to get back to sleep”. Sleepiness was assessed by the ESS as well as by self-report of being excessively sleepy during the day most or almost all of the time.

### Statistical Analyses

Mean and standard deviation, and percentages were used to provide an overall description of the data used in the analyses. Unadjusted differences were compared using Student’s t test or χ^2^. For both definitions of the AHI, each participant’s AHI was assigned to one of 4 OSA severity categories: Normal (AHI <5 /hour), Mild (AHI ≥5 and <15 /hour), Moderate (AHI ≥15 and < 30/hour) and Severe (AHI ≥30/hour).

Missing data was present in 4.8% of observations and were felt to be missing at random; multiple imputation using the mice package in R was employed to generate replacement values. Comparison of the imputed to the original dataset did not identify any outliers in the imputed dataset and means of the same variables between datasets were comparable.

To reduce overfitting of models and reduce potential collinearity, a Least Absolute Shrinkage and Selection Operator (lasso) regression was performed for both outcome variables using the glmnet package in R. This resulted in an analytic dataset for CVD that consisted of the following: age, sex, race/ethnicity, BMI, AAI, diastolic BP, smoking, SF-36 physical component summary (PCS), SF-36 general health rating (GenHlth), SF-36 ability to perform vigorous activity (VigActiv), hypertension, diabetes, depression and HDL. For CHD, the analytic dataset consisted of the following: age, sex, race/ethnicity, BMI, diastolic BP, smoking, PCS, GenHlth, VigActiv, hypertension, diabetes, triglycerides and HDL.

For the entire cohort, logistic regression using SPSS v27 (Armonk, NY) was used to generate increasingly complex models of the relationship between either CVD or CHD and severity of OSA adjusting for the covariates identified using the lasso regression. For CVD, after the unadjusted model, models were generated for the sequential addition of demographic factors (age, sex, race/ethnicity), anthropometric factors (BMI, AAI, diastolic BP), social/behavioral characteristics (smoking, PCS, GenHlth, VigActiv) and diseases/conditions (hypertension, diabetes, depression, HDL). For CHD after the unadjusted model, the corresponding sequential models were demographic factors (age, sex, race/ethnicity), anthropometric factors (BMI, diastolic BP), social/behavioral characteristics (smoking, PCS, GenHlth, VigActiv) and diseases/conditions (hypertension, diabetes, triglycerides, HDL). Because of the possibility that adjustment for a hypertension and a diabetes indicator would be “overadjustment” (i.e., adjustment for a variable on a causal pathway), we excluded both hypertension and diabetes from the final set of covariates in additional analyses and these are referred to as “parsimonious models.” Lastly, sensitivity analyses were performed in which the natural log of AHI3%A was used instead of categorial levels of that factor in the above models.

Associations between both CVD and CHD, and OSA severity were further analyzed in the subgroup of participants who were not classified as having OSA based on AHI4% criteria but were classified as OSA using AHI3%A criteria. The moderate and severe categories were combined because of the small number of cases in the severe OSA category. Otherwise, the modelling approaches employed were identical.

In Tables 2-5, odds ratios and 95% CI are presented versus the reference level of AHI <5 /hour. P values refer to the overall significance of the model with respect to OSA severity. Odds ratios, 95% CI and P values in Table 6 refer to AHI3%A expressed as the continuous factor lnAHI3%+0.1 (0.1 added to mitigate 0 values of lnAHI3%).

**Table 1:**
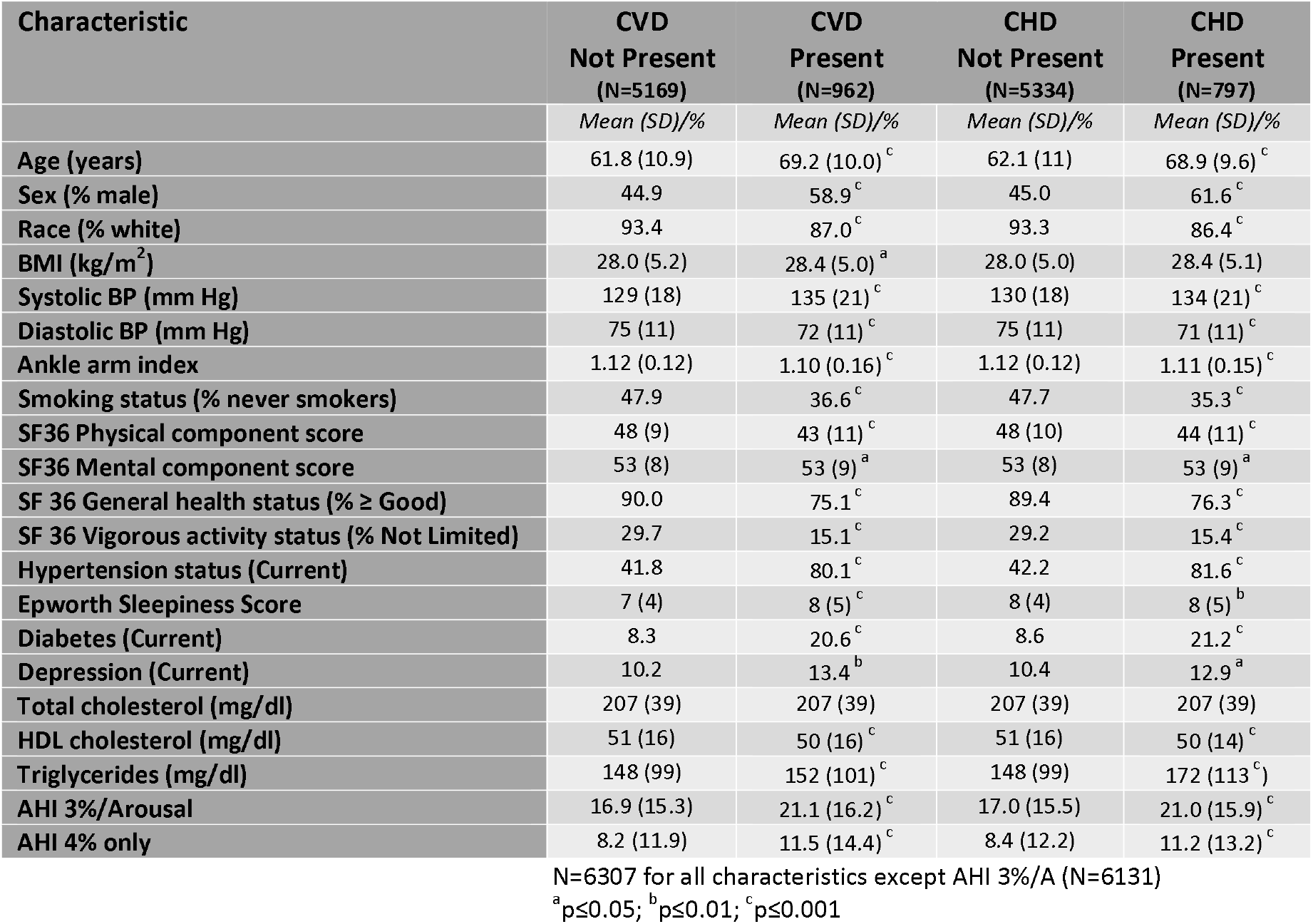
Univariate Association of Various Characteristics to Prevalent Cardiovascular (CVD) and Coronary Heart Disease (CHD)

| Characteristic | CVD<br>Not Present<br>(N=5169) | CVD<br>Present<br>(N=962) | CHD<br>Not Present<br>(N=5334) | CHD<br>Present<br>(N=797) |
| --- | --- | --- | --- | --- |
|  | Mean (SD)/% | Mean (SD)/% | Mean (SD)/% | Mean (SD)/% |
| Age (years) | 61.8 (10.9) | 69.2 (10.0)<sup>c</sup> | 62.1 (11) | 68.9 (9.6)<sup>c</sup> |
| Sex (% male) | 44.9 | 58.9<sup>c</sup> | 45.0 | 61.6<sup>c</sup> |
| Race (% white) | 93.4 | 87.0<sup>c</sup> | 93.3 | 86.4<sup>c</sup> |
| BMI (kg/m<sup>2</sup>) | 28.0 (5.2) | 28.4 (5.0)<sup>a</sup> | 28.0 (5.0) | 28.4 (5.1) |
| Systolic BP (mm Hg) | 129 (18) | 135 (21)<sup>c</sup> | 130 (18) | 134 (21)<sup>c</sup> |
| Diastolic BP (mm Hg) | 75 (11) | 72 (11)<sup>c</sup> | 75 (11) | 71 (11)<sup>c</sup> |
| Ankle arm index | 1.12 (0.12) | 1.10 (0.16)<sup>c</sup> | 1.12 (0.12) | 1.11 (0.15)<sup>c</sup> |
| Smoking status (% never smokers) | 47.9 | 36.6<sup>c</sup> | 47.7 | 35.3<sup>c</sup> |
| SF36 Physical component score | 48 (9) | 43 (11)<sup>c</sup> | 48 (10) | 44 (11)<sup>c</sup> |
| SF36 Mental component score | 53 (8) | 53 (9)<sup>a</sup> | 53 (8) | 53 (9)<sup>a</sup> |
| SF 36 General health status (% ≥ Good) | 90.0 | 75.1<sup>c</sup> | 89.4 | 76.3<sup>c</sup> |
| SF 36 Vigorous activity status (% Not Limited) | 29.7 | 15.1<sup>c</sup> | 29.2 | 15.4<sup>c</sup> |
| Hypertension status (Current) | 41.8 | 80.1<sup>c</sup> | 42.2 | 81.6<sup>c</sup> |
| Epworth Sleepiness Score | 7 (4) | 8 (5)<sup>c</sup> | 8 (4) | 8 (5)<sup>b</sup> |
| Diabetes (Current) | 8.3 | 20.6<sup>c</sup> | 8.6 | 21.2<sup>c</sup> |
| Depression (Current) | 10.2 | 13.4<sup>b</sup> | 10.4 | 12.9<sup>a</sup> |
| Total cholesterol (mg/dl) | 207 (39) | 207 (39) | 207 (39) | 207 (39) |
| HDL cholesterol (mg/dl) | 51 (16) | 50 (16)<sup>c</sup> | 51 (16) | 50 (14)<sup>c</sup> |
| Triglycerides (mg/dl) | 148 (99) | 152 (101)<sup>c</sup> | 148 (99) | 172 (113)<sup>c</sup> |
| AHI 3%/Arousal | 16.9 (15.3) | 21.1 (16.2)<sup>c</sup> | 17.0 (15.5) | 21.0 (15.9)<sup>c</sup> |
| AHI 4% only | 8.2 (11.9) | 11.5 (14.4)<sup>c</sup> | 8.4 (12.2) | 11.2 (13.2)<sup>c</sup> |

N=6307 for all characteristics except AHI 3%/A (N=6131)
<sup>a</sup>p≤0.05; <sup>b</sup>p≤0.01; <sup>c</sup>p≤0.001

**Table 2.**
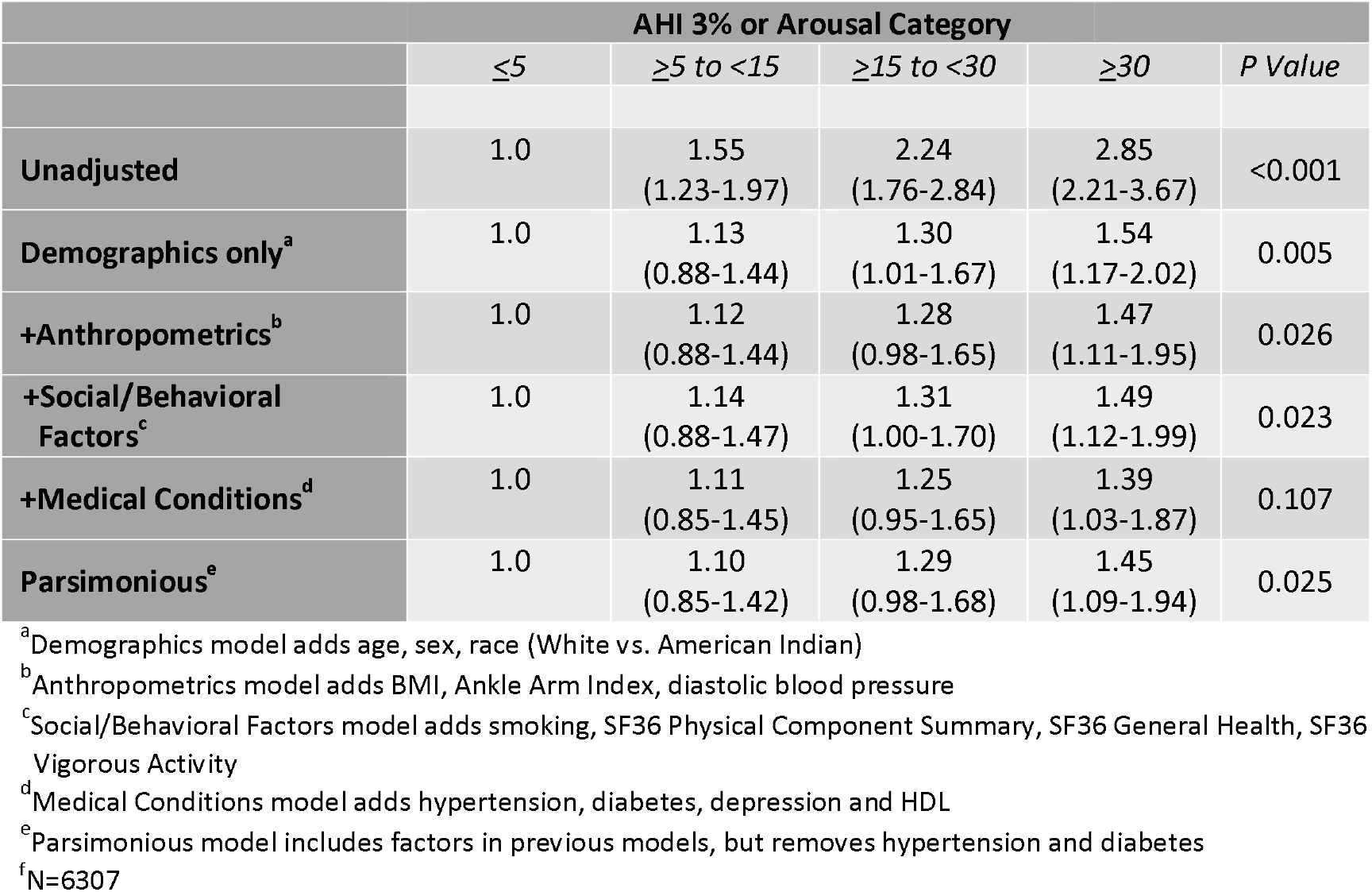
Adjusted Relative Odds (95% Confidence Interval) of Self-Reported Prevalent Cardiovascular Disease According to 3% or Arousal Apnea Hypopnea Index Severity Categories.

|  | **AHI 3% or Arousal Category** |  |  |  | *P Value* |
| --- | --- | --- | --- | --- | --- |
| | $\leq 5$ | $\geq 5$ to $<15$ | $\geq 15$ to $<30$ | $\geq 30$ | |
| **Unadjusted** | 1.0 | 1.55<br>(1.23-1.97) | 2.24<br>(1.76-2.84) | 2.85<br>(2.21-3.67) | <0.001 |
| **Demographics only<sup>a</sup>** | 1.0 | 1.13<br>(0.88-1.44) | 1.30<br>(1.01-1.67) | 1.54<br>(1.17-2.02) | 0.005 |
| **+Anthropometrics<sup>b</sup>** | 1.0 | 1.12<br>(0.88-1.44) | 1.28<br>(0.98-1.65) | 1.47<br>(1.11-1.95) | 0.026 |
| **+Social/Behavioral Factors<sup>c</sup>** | 1.0 | 1.14<br>(0.88-1.47) | 1.31<br>(1.00-1.70) | 1.49<br>(1.12-1.99) | 0.023 |
| **+Medical Conditions<sup>d</sup>** | 1.0 | 1.11<br>(0.85-1.45) | 1.25<br>(0.95-1.65) | 1.39<br>(1.03-1.87) | 0.107 |
| **Parsimonious<sup>e</sup>** | 1.0 | 1.10<br>(0.85-1.42) | 1.29<br>(0.98-1.68) | 1.45<br>(1.09-1.94) | 0.025 |

<sup>a</sup>Demographics model adds age, sex, race (White vs. American Indian)
<sup>b</sup>Anthropometrics model adds BMI, Ankle Arm Index, diastolic blood pressure
<sup>c</sup>Social/Behavioral Factors model adds smoking, SF36 Physical Component Summary, SF36 General Health, SF36 Vigorous Activity
<sup>d</sup>Medical Conditions model adds hypertension, diabetes, depression and HDL
<sup>e</sup>Parsimonious model includes factors in previous models, but removes hypertension and diabetes
<sup>f</sup>N=6307

**Table 3.** Adjusted Relative Odds (95% Confidence Interval) of Self-Reported Prevalent Coronary Heart Disease According to 3% or Arousal Apnea Hypopnea Index Severity Categoriesf.

| AHI 3% or Arousal Category |  |  |  |  |  |
| --- | --- | --- | --- | --- | --- |
|  | <5 | ≥5 to <15 | ≥15 to <30 | ≥30 | P Value |
| **Unadjusted** | 1.0 | 1.57<br>(1.21-2.04) | 2.38<br>(1.83-3.10) | 2.81<br>(2.12-3.71) | <0.001 |
| **Demographics only** | 1.0 | 1.14<br>(0.87-1.49) | 1.38<br>(1.05-1.82) | 1.48<br>(1.10-1.99) | 0.016 |
| **+Anthropometrics** | 1.0 | 1.13<br>(0.86-1.49) | 1.34<br>(1.01-1.77) | 1.40<br>(1.03-1.90) | 0.07 |
| **+Social/Behavioral Factors** | 1.0 | 1.14<br>(0.87-1.51) | 1.37<br>(1.03-1.82) | 1.42<br>(1.04-1.94) | 0.053 |
| **+Medical Conditions** | 1.0 | 1.11<br>(0.83-1.47) | 1.29<br>(0.96-1.74) | 1.29<br>(0.96-1.74) | 0.222 |
| **Parsimonious** | 1.0 | 1.08<br>(0.82-1.43) | 1.31<br>(0.99-1.75) | 1.36<br>(0.99-1.85) | 0.073 |

<sup>a</sup>Demographics model adds age, sex, race (White vs. American Indian)
<sup>b</sup>Anthropometrics model adds BMI, diastolic blood pressure
<sup>c</sup>Social/Behavioral Factors model adds smoking, SF36 Physical Component Summary, SF36 General Health, SF36 Vigorous Activity
<sup>d</sup>Medical Conditions model adds hypertension, diabetes, triglycerides and HDL
<sup>e</sup>Parsimonious model includes factors in previous models, but removes hypertension and diabetes
<sup>f</sup>N=6307

**Table 4.** Adjusted Relative Odds (95% Confidence Interval) of Self-Reported Prevalent Cardiovascular Disease According to 3% or Arousal Apnea Hypopnea Index Severity Categories in Participants Without Obstructive Sleep Apnea According to 4% Desaturation Criteria^f^.

| **AHI 3% or Arousal Category** |  |  |  |  |
| --- | --- | --- | --- | --- |
|  | **≤5** | **>5 to ≤15** | **>15** | **P Value** |
| **Unadjusted** | 1.0 | 1.42<br>(1.11-1.81) | 2.55<br>(1.79-3.63) | <0.001 |
| **Demographics only<sup>a</sup>** | 1.0 | 1.02<br>(0.79-1.29) | 1.46<br>(1.00-2.14) | 0.091 |
| **+Anthropometrics<sup>b</sup>** | 1.0 | 0.99<br>(0.77-1.30) | 1.49<br>(1.02-2.18) | 0.058 |
| **+Social/Behavioral Factors<sup>c</sup>** | 1.0 | 1.01<br>(0.78-1.32) | 1.53<br>(1.04-2.26) | 0.053 |
| **+Medical Conditions<sup>d</sup>** | 1.0 | 0.98<br>(0.75-1.30) | 1.50<br>(1.00-2.26) | 0.067 |
| **Parsimonious<sup>e</sup>** | 1.0 | 0.97<br>(0.74-1.27) | 1.47<br>(1.00-2.18) | 0.056 |

<sup>a</sup>Demographics model adds age, sex, race (White vs. American Indian)
<sup>b</sup>Anthropometrics model adds BMI, Ankle Arm Index, diastolic blood pressure
<sup>c</sup>Social/Behavioral Factors model adds smoking, SF36 Physical Component Summary, SF36 General Health, SF36 Vigorous Activity
<sup>d</sup>Medical Conditions model adds hypertension, diabetes, depression and HDL
<sup>e</sup>Parsimonious model includes factors in previous models, but removes hypertension and diabetes
<sup>f</sup>N=3326

**Table 5.** Adjusted Relative Odds (95% Confidence Interval) of Self-Reported Prevalent Coronary Heart Disease According to 3% or Arousal Apnea Hypopnea Index Severity Categories in Participants Without Obstructive Sleep Apnea According to 4% Desaturation Criterion^f^.

| **AHI 3% or Arousal Category** |  |  |  |  |
| --- | --- | --- | --- | --- |
|  | **≤5** | **>5 to ≤15** | **>15** | **P Value** |
| **Unadjusted** | 1.0 | 1.45<br>(1.11-1.90) | 2.70<br>(1.85-3.96) | <0.001 |
| **Demographics only<sup>a</sup>** | 1.0 | 1.06<br>(0.80-1.40) | 1.57<br>(1.05-2.35) | 0.061 |
| **+Anthropometrics<sup>b</sup>** | 1.0 | 1.02<br>(0.77-1.36) | 1.53<br>(1.02-2.30) | 0.066 |
| **+Social/Behavioral Factors<sup>c</sup>** | 1.0 | 1.04<br>(0.78-1.38) | 1.58<br>(1.05-2.38) | 0.056 |
| **+Medical Conditions<sup>d</sup>** | 1.0 | 1.02<br>(0.76-1.37) | 1.57<br>(1.02-2.41) | 0.066 |
| **Parsimonious<sup>e</sup>** | 1.0 | 0.99<br>(0.74-1.32) | 1.52<br>(1.01-2.30) | 0.058 |

<sup>a</sup>Demographics model adds age, sex, race (White vs. American Indian)
<sup>b</sup>Anthropometrics model adds BMI, diastolic blood pressure
<sup>c</sup>Social/Behavioral Factors model adds smoking, SF36 Physical Component Summary, SF36 General Health, SF36 Vigorous Activity
<sup>d</sup>Medical Conditions model adds hypertension, diabetes, triglycerides and HDL
<sup>e</sup>Parsimonious model includes factors in previous models, but removes hypertension and diabetes
<sup>f</sup>N=3326

**Table 6.** Linear Adjusted Relative Odds (95% Confidence Interval) of Self-Reported Prevalent Cardiovascular and Coronary Heart Disease According to 3% or Arousal Apnea Hypopnea Index Severity.

|  | Odds Ratio<br>(95% CI) | P Value | Odds Ratio<br>(95% CI) | P Value |
| --- | --- | --- | --- | --- |
| **Full Cohort** | **CVD** |  | **CHD** |  |
| **Unadjusted** | 1.46<br>(1.35-1.58) | <0.001 | 1.45<br>(1.33-1.58) | <0.001 |
| **Fully Adjusted<sup>b</sup>** | 1.07<br>(0.98-1.17) | 0.152 | 1.10<br>(1.00-1.22) | 0.060 |
| **Parsimonious<sup>c</sup>** | 1.12<br>(1.03-1.23) | 0.010 | 1.13<br>(1.02-1.25) | 0.016 |
| **Restricted Cohort<sup>a</sup>** | **CVD** |  | **CHD** |  |
| **Unadjusted** | 1.57<br>(1.34-1.84) | <0.001 | 1.62<br>(1.36-1.93) | <0.001 |
| **Fully Adjusted<sup>b</sup>** | 1.18<br>(1.00-1.40) | 0.055 | 1.22<br>(1.01-1.47) | 0.036 |
| **Parsimonious<sup>c</sup>** | 1.17<br>(0.99-1.38) | 0.072 | 1.20<br>(1.00-1.44) | 0.055 |

<sup>a</sup>Cohort restricted participants without OSA according to AHI4% criteria, N=3326
<sup>b</sup>Covariates for CVD: age, sex, race, BMI, ankle-arm index, diastolic blood pressure, smoking, SF36 Physical Component Summary, SF36 General Health, SF36 Vigorous Activity, hypertension, diabetes, depression and HDL; Covariates for CHD: age, sex, race, BMI, diastolic blood pressure, smoking, SF36 Physical Component Summary, SF36 General Health, SF36 Vigorous Activity, hypertension, diabetes, triglycerides and HDL
<sup>c</sup>Excludes diabetes and hypertension from fully adjusted model

## Results

Table 1 shows the univariate association of potential risk factors or characteristics with the presence of CVD or CHD. There were 962 cases (15%) of CVD and 797 (13%) cases of CHD identified. Except for total cholesterol, all were either more prevalent or significantly higher or lower in participants with CVD or CHD. For both CVD and CHD, markedly higher prevalence rates were noted for sex (higher in men), hypertension, diabetes, depression, smoking (higher in ever smokers) and ability to engage in vigorous activity. Conversely, white race and good health status were much less common among those with CVD or CHD. Differences observed for the remaining characteristics were of lesser magnitude.

In Figure 1 shows the prevalence rates of CVD or CHD as a function of OSA severity using the AHI3%A criteria. Both conditions were associated with increasing higher rates of disease as OSA became more severe.

**Figure:**
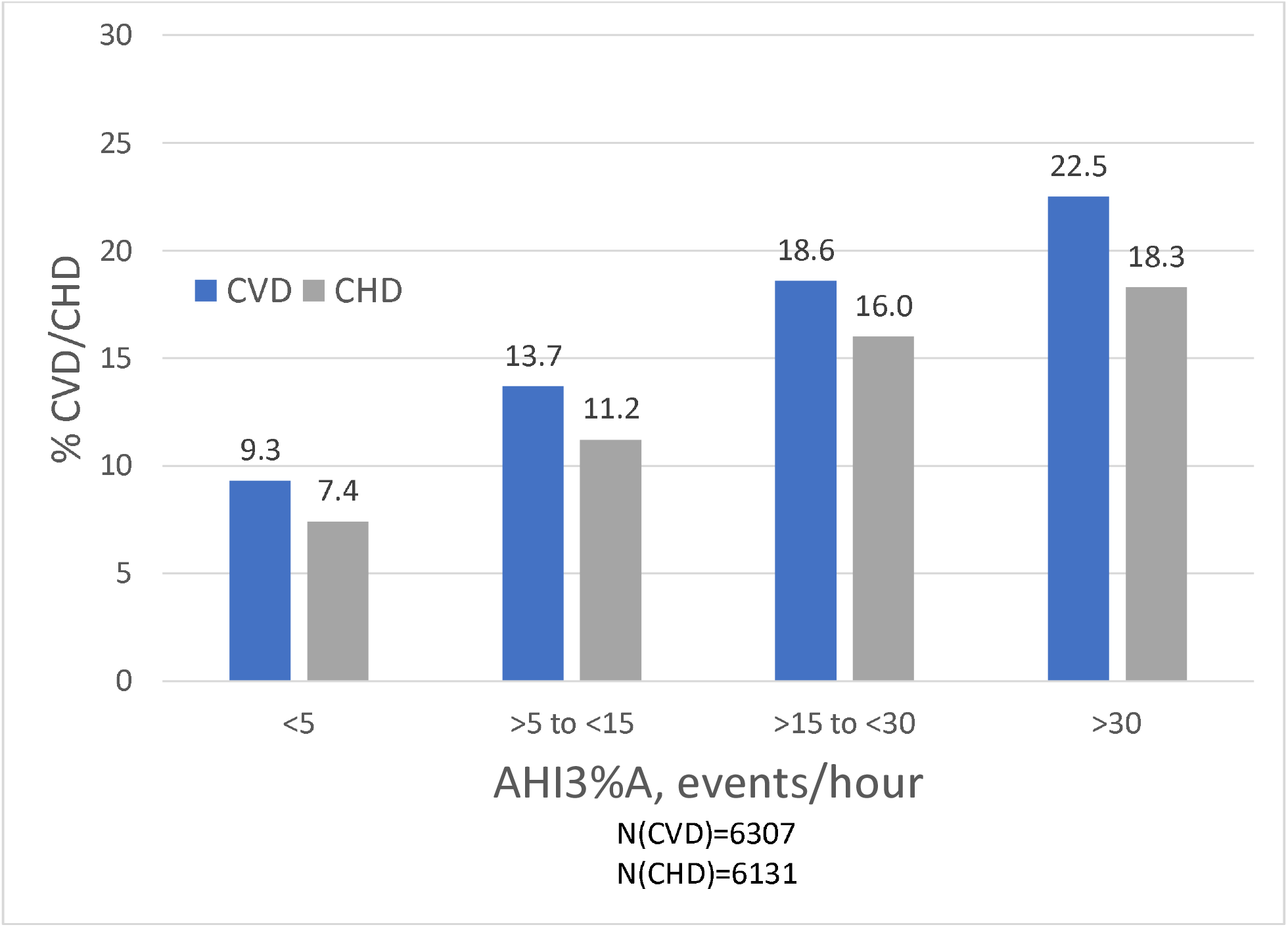
Percentage of participants with either cardiovascular (CVD) or coronary heart (CHD) disease according to increasing severity of obstructive sleep apnea defined using a hypopnea definition characterized by a minimum 3% oxygen desaturation or an arousal (AHI3%A)

Table 2 shows the crude and adjusted odds ratios and their 95% confidence intervals for increasing complex models of the relationship between CVD and AHI3%A. The unadjusted model showed a strong, progressive association with increasingly severe OSA. However, as the models became increasingly complex, this relationship was attenuated and only approached statistical significance in the fully adjusted model (+Medical Conditions). Removal of hypertension and diabetes to create the Parsimonious model restored some of the association with a return of statistical significance.

Presented in Table 3 are the models demonstrating the relationship between CHD and AHI3%A. Similar to the findings for CVD, there was a progressively higher odds of having CHD as severity of OSA increased. The fully adjusted model (+Medical Conditions) was not significant, but the Parsimonious model approached statistical significance.

There were 3,326 participants who did not have OSA as defined by AHI4% criteria. Within this cohort, 2247 were classified as OSA using the AHI3%A criteria; 1966 (87.4%) were mild, 271 (12.0%) were moderate and 10 (0.4%) were severe. For this subgroup, Table 4 presents the increasingly complex models illustrating the relationship between the presence of CVD and increasing OSA severity. Because of the relatively small number of cases with severe OSA, the moderate and severe cases were combined for these analyses. The unadjusted model showed a strong relationship with OSA severity; as model complexity increased, this finding was attenuated and only approached statistical significance in both the fully adjusted (+Medical Conditions) and Parsimonious models. As demonstrated in Table 5, similar findings were observed for CHD; the unadjusted model showed a strong association which was attenuated as the models became more complex; the fully adjusted (+medical conditions) and parsimonious models approached statistical significance.

In sensitivity analyses, the natural log of AHI3%A was used as the index of OSA severity in lieu of a categorial representation. As shown in Table 6, in the entire cohort, for both CVD and CHD, a significant linear relationship with increasing severity of OSA was demonstrated in parsimonious models, but not the fully adjusted models. In the subgroup who did not have OSA as defined by AHI4% criteria but did have OSA using the AHI3%A criteria, linear relationships noted for both CVD and CHD in the fully adjusted and parsimonious models. For CHD in the fully adjusted model, the relationship was statistically significant and approached statistical significance in the others.

## Discussion

In this large community-based study, we demonstrated that OSA defined by apneas and hypopneas characterized by 3% desaturation events or arousals is associated with an increased likelihood of self-reported CVD and CHD after controlling for a number of relevant covariates. Importantly, in a subset of this cohort who did not have OSA as defined by apneas and hypopneas requiring a minimum 4% oxygen desaturation but did have OSA using the 3% desaturation or arousal criteria, we found that the association with both CVD and CHD remained, albeit weaker. Nevertheless, our analyses overall suggest that the regulatory requirement by the Centers for Medicare and Medicaid Services (CMS) in the United States of using a 4% desaturation definition to identify patients with OSA denies a substantial proportion of these individuals the opportunity to be treated for their OSA and thus reduce the risk of worsening or recurrence of their CVD or CHD.

Results from several large cohort studies including SHHS have found that OSA is associated with the presence of CVD and CHD, and that this association is stronger when the AHI as a metric of OSA severity increases (1, 2). These previous studies have used a definition of hypopnea that requires a minimum 4% oxygen desaturation (16-18). This definition has been adopted by CMS and other insurers to identify individuals as having OSA (5). However, the AASM recommends defining hypopneas with a minimum 3% desaturation or an arousal (4). This is based on evidence indicating that daytime sleepiness and other symptoms of OSA are associated with this less stringent definition of OSA (6). This distinction has important clinical implications because there are a large number of patients who do not meet the AHI4% criteria and but do meet the AHI3%A criteria (7, 19). In the former case, they are not considered to have OSA, but do have it in the latter.

To our knowledge, our study is the first to assess the association between OSA using the AHI3%A criteria and CVD and CHD. We found that as OSA severity increased, there was a greater likelihood of having CVD and CHD after adjusting for a number of relevant covariates. We acknowledge that in the fully adjusted model, this association only approached statistical significance. However, in our parsimonious model which removed the presence of hypertension and diabetes, likely mediators of this relationship, the association was strengthened. Sensitivity analyses using the natural log of AHI3%A validated the results we observed with categories of AHI severity. It has been well-established that hypertension and diabetes are independent risk factors for the development of CVD and CHD. However, a number of studies have demonstrated that OSA is a risk factor for the development of both hypertension and diabetes (1, 20). Therefore, both of the latter conditions lie on the causal path by which OSA may increase the risk for the development of CVD and CHD. Hence, we believe that inclusion of both these conditions in our fully adjusted model may be over- adjustment and that our parsimonious model best represents the association between OSA defined by AHI3%A and CVD or CHD.

We identified there was a large subset of our cohort that had OSA using the AHI3%A definition, but not the AHI4% definition of hypopnea. In this subset, we also observed an association between OSA severity and both CVD and CHD. This finding is analogous to the relationship we recently observed between OSA and the prevalence of hypertension (21). Similar to our findings with the full cohort, the fully adjusted model for both CVD and CHD was not statistically significant. However, it approached or became statistically significant in the parsimonious models. Although most of these cases were in the mild OSA category, 12.4% were moderate to severe where treatment is almost always recommended. Individuals with prevalent CVD or CHD and OSA are at risk for further complications of their CVD or CHD (22-24). However, if they do not meet the AHI4% definition of OSA, access to OSA treatment would be denied by CMS and some insurers.

Our findings with respect to CVD and CHD are consistent with recent analyses demonstrating that OSA defined by AHI3%A is associated with prevalent and incident hypertension in the SHHS cohort (19, 25). Similar findings also have been observed in other cohorts providing additional evidence that use of a hypopnea definition incorporating a minimum 3% oxygen desaturation or an arousal is important in the identification of individuals with OSA (26-28).

Although there is substantial evidence emerging that intermittent hypoxemia plays an important role in the cardiovascular consequences of OSA (29), the importance of arousals remains uncertain (30). Arousals involve an increase in the sympathetic activity and a decrease in the parasympathetic activity (29) and there is some evidence linking them in the development of hypertension (31). Data from our study would suggest that they may contribute to the development of CVD or CHD as well.

Some (32, 33), but not all (34) studies have suggested that the impact of OSA on the development of CVD or CHD is in part enhanced by the presence of sleepiness. However, in our initial assessment of potential covariates using a lasso regression, sleepiness did not emerge as a significant factor. Thus, our findings do not support the contention that sleepiness is an important factor impacting the relationship between OSA and CVD or CHD.

Our study does have a few limitations. Most important is that we identified prevalent CVD and CHD by self-report. While it is possible that some misclassification occurred, we do not think it was large. A large number of potential covariates were considered for inclusion in the models; we used a lasso regression to reduce the possibility of over-adjustment and collinearity. Furthermore, the possibility of residual confounding remains. Finally, this is a cross-sectional analysis, and causality cannot be assumed.

This study has several strengths. It uses a large, well characterized cohort with the availability of data from a number of potential covariates. Additionally, the cohort had a diverse racial/ethnic, age and sex distribution. Polysomnography was used to document the presence of OSA, and not more limited sleep apnea testing.

In summary, OSA as defined by apneas and hypopneas requiring a minimum 3% oxygen desaturation or arousal is associated with an increased likelihood of having CVD or CHD. Use of a more restrictive definition requiring a minimum 4% desaturation will misidentify a large number of individuals with OSA, and CVD or CHD. These individuals may be denied access to therapy which may prevent worsening of their underlying CVD or CHD.

## Data Availability

SHHS data is available from the Dr. Quan or through the website: https://sleepdata.org/datasets/shhs

## Acknowledgements

SHHS acknowledges the Atherosclerosis Risk in Communities Study, the Cardiovascular Health Study, the Framingham Heart Study, the Cornell/Mt. Sinai Worksite and Hypertension Studies, the Strong Heart Study, the Tucson Epidemiologic Study of Airways Obstructive Diseases (TESAOD), and the Tucson Health and Environment Study for allowing their cohort members to be part of the SHHS and for sharing such data for the purposes of this study. SHHS is particularly grateful to the members of these cohorts who agreed to participate in SHHS as well. SHHS further recognizes all the investigators and staff who have contributed to its success. A list of SHHS investigators, staff, and their participating institutions is available on the SHHS website (www.jhsph.edu/shhs).

The opinions expressed in this paper are those of the authors and do not necessarily reflect the views of the Indian Health Service.

This work was supported by National Heart, Lung and Blood Institute cooperative agreements U01HL53940 (University of Washington), U01HL53941 (Boston University), U01HL53938 (University of Arizona), U01HL53916 (University of California, Davis), U01HL53934 (University of Minnesota), U01HL53931 (New York University), U01HL53937 and U01HL64360 (Johns Hopkins University), U01HL63463 (Case Western Reserve University), U01HL63429 (Missouri Breaks Research).

SP was supported by Patient Centered Outcomes Research Institute (CER- 2018C2-13262; PCS-1504-30430; DI-2018C2-13161; DI-2018C2-13161 COVID supplement, EADI-16493), NIH (HL126140, HL151254, AI135108, AG059202, HL158253) and American Academy of Sleep Medicine Foundation during the writing of this manuscript.

## Abbreviation List

AHI: Apnea Hypopnea Index
AHI3%A: Hypopneas with at least a 3% oxygen desaturation or an arousal
AHI4%: Hypopneas with at least a 4% oxygen desaturation
AAI: Ankle arm index
BP: Blood pressure
BMI: Body mass index
CHD: Coronary heart disease
CVD: Cardiovascular disease
CMS: Centers for Medicare and Medicaid Services
ESS: Epworth sleepiness scale
GenHlth: General health rating subscale of SF36
HDL: High density lipoprotein
Lasso: Least Absolute Shrinkage and Selection Operator
OSA: Obstructive sleep apnea
PCS: Physical component summary of the SF36
SHHS: Sleep Heart Health Study
VigActiv: Vigorous activity rating subscale of the SF36

## References

1. Hou H, Zhao Y, Yu W, et al. Association of obstructive sleep apnea with hypertension: A systematic review and meta-analysis. J Glob Health. 2018;8(1):010405. 10.7189/jogh.08.010405 [doi] PMID: 29497502

2. Zhang X, Fan J, Guo Y, et al. Association between obstructive sleep apnoea syndrome and the risk of cardiovascular diseases: an updated systematic review and dose-response meta-analysis. Sleep Med. 2020;7139–46. 10.1016/j.sleep.2020.03.011 PMID: 29497502

3. Mashaqi S, Gozal D. The impact of obstructive sleep apnea and PAP therapy on all-cause and cardiovascular mortality based on age and gender -a literature review. Respir Investig. 2020;58(1):7–20. S2212-5345(19)30159-5 [pii] PMID: 31631059

4. Berry RB, Budhiraja R, Gottlieb DJ, et al. Rules for scoring respiratory events in sleep: update of the 2007 AASM Manual for the Scoring of Sleep and Associated Events. Deliberations of the Sleep Apnea Definitions Task Force of the American Academy of Sleep Medicine. J Clin Sleep Med. 2012;8(5):597-619. 10.5664/jcsm.2172 [doi] PMID: 23066376

5. Anonymous. CPAP for Obstructive Sleep Apnea. Last Updated: 2020. Accessed: 09/14, 2020. https://www.cms.gov/Medicare/Coverage/Coverage-with-Evidence-Development/CPAP.

6. Shamin-Uzzaman QA, Singh S, Chowdhuri S. Hypopnea definitions, determinants, and dilemmas: a focused review. Sci Sleep Pract. 2018;2(1):7.

7. Raj R, Hirshkowitz M. Effect of the new Medicare guideline on patient qualification for positive airway pressure therapy. Sleep Med. 2003;4(1):29–33. S1389945702001508 [pii] PMID: 14592357

8. Quan SF, Howard BV, Iber C, et al. The Sleep Heart Health Study: design, rationale, and methods. Sleep. 1997;20(12):1077–1085. PMID: 9493915

9. Lind BK, Goodwin JL, Hill JG, Ali T, Redline S, Quan SF. Recruitment of healthy adults into a study of overnight sleep monitoring in the home: experience of the Sleep Heart Health Study. Sleep Breath. 2003;7(1):13-24. 10.1007/s11325-003-0013-z PMID: 12712393

10. Iber C, Redline S, Kaplan Gilpin AM, et al. Polysomnography performed in the unattended home versus the attended laboratory setting--Sleep Heart Health Study methodology. Sleep. 2004;27(3):536–540. PMID: 15164911

11. Johns MW. A new method for measuring daytime sleepiness: the Epworth sleepiness scale. Sleep. 1991;14(6):540–545. PMID: 1798888

12. Ware JE,Jr, Sherbourne CD. The MOS 36-item short-form health survey (SF-36). I. Conceptual framework and item selection. Med Care. 1992;30(6):473–483. PMID: 1593914

13. O’Connor GT, Caffo B, Newman AB, et al. Prospective study of sleep-disordered breathing and hypertension: the Sleep Heart Health Study. Am J Respir Crit Care Med. 2009;179(12):1159-1164. 10.1164/rccm.200712-1809OC PMID: 19264976; 200712-1809OC [pii]

14. Whitney CW, Gottlieb DJ, Redline S, et al. Reliability of scoring respiratory disturbance indices and sleep staging. Sleep. 1998;21(7):749-757. 10.1093/sleep/21.7.749 [doi] PMID: 11286351

15. Rechtschaffen A, Kales A. A Manual of Standardized Terminology, Techniques and Scoring System for Sleep Stages of Human Subject. Washington, D.C.: US Government Printing Office, National Institute of Health Publication, 1968;

16. Marin JM, Carrizo SJ, Vicente E, Agusti AG. Long-term cardiovascular outcomes in men with obstructive sleep apnoea-hypopnoea with or without treatment with continuous positive airway pressure: an observational study. Lancet. 2005;365(9464):1046–1053. 10.1016/S0140-6736(05)71141-7 PMID: 15781100; S0140-6736(05)71141-7 [pii]

17. Shahar E, Whitney CW, Redline S, et al. Sleep-disordered breathing and cardiovascular disease: cross-sectional results of the Sleep Heart Health Study. Am J Respir Crit Care Med. 2001;163(1):19-25. 11208620

18. Young T, Finn L, Peppard PE, et al. Sleep disordered breathing and mortality: eighteen-year follow-up of the Wisconsin sleep cohort. Sleep. 2008;31(8):1071–1078. PMID: 18714778

19. Budhiraja R, Javaheri S, Parthasarathy S, Berry RB, Quan SF. The Association Between Obstructive Sleep Apnea Characterized by a Minimum 3 Percent Oxygen Desaturation or Arousal Hypopnea Definition and Hypertension. J Clin Sleep Med. 2019;15(9):1261-1270. 10.5664/jcsm.7916 [doi] PMID: 31538597

20. Qie R, Zhang D, Liu L, et al. Obstructive sleep apnea and risk of type 2 diabetes mellitus: A systematic review and dose-response meta-analysis of cohort studies. J Diabetes. 2020;12(6):455–464. 10.1111/1753-0407.13017 [doi] PMID: 31872550

21. Budhiraja R, Javaheri S, Parthasarathy S, Berry RB, Quan SF. The Association Between Obstructive Sleep Apnea Characterized by a Minimum 3 Percent Oxygen Desaturation or Arousal Hypopnea Definition and Hypertension. J Clin Sleep Med. 2019;15(9):1261–1270. 10.5664/jcsm.7916 [doi] PMID: 31538597

22. Yudi MB, Clark DJ, Farouque O, et al. Trends and predictors of recurrent acute coronary syndrome hospitalizations and unplanned revascularization after index acute myocardial infarction treated with percutaneous coronary intervention. Am Heart J. 2019;212134-143. S0002-8703(19)30048-1 [pii] PMID: 31004916

23. Lee CH, Khoo SM, Chan MY, et al. Severe obstructive sleep apnea and outcomes following myocardial infarction. J Clin Sleep Med. 2011;7(6):616–621. 10.5664/jcsm.1464 [doi] PMID: 22171200

24. Maia FC, Goulart AC, Drager LF, et al. Impact of High Risk for Obstructive Sleep Apnea on Survival after Acute Coronary Syndrome: Insights from the ERICO Registry. Arq Bras Cardiol. 2017;108(1):31–37. S0066-782X2017000100031 [pii] PMID: 28146212

25. Budhiraja R, Javaheri S, Parthasarathy S, Berry RB, Quan SF. Incidence of hypertension in obstructive sleep apnea using hypopneas defined by a 3 percent oxygen desaturation or arousal but not by only 4 percent oxygen desaturation. J Clin Sleep Med. 2020;10.5664/jcsm.8684 [doi] PMID: 32643602

26. Bouloukaki I, Grote L, McNicholas WT, et al. Mild obstructive sleep apnea increases hypertension risk, challenging traditional severity classification. J Clin Sleep Med. 2020;16(6):889–898. 10.5664/jcsm.8354 [doi] PMID: 32043960

27. Hirotsu C, Haba-Rubio J, Andries D, et al. Effect of Three Hypopnea Scoring Criteria on OSA Prevalence and Associated Comorbidities in the General Population. J Clin Sleep Med. 2019;15(2):183–194. 10.5664/jcsm.7612 [doi] PMID: 30736872

28. Heinzer R, Vat S, Marques-Vidal P, et al. Prevalence of sleep-disordered breathing in the general population: the HypnoLaus study. Lancet Respir Med. 2015;3(4):310–318. 10.1016/S2213-2600(15)00043-0 [doi] PMID: 25682233

29. Sforza E, Roche F. Chronic intermittent hypoxia and obstructive sleep apnea: an experimental and clinical approach. Hypoxia (Auckl). 2016;499–108. hp-4-099 [pii] PMID: 27800512

30. Ryan S. Mechanisms of cardiovascular disease in obstructive sleep apnoea. J Thorac Dis. 2018;10(Suppl 34):S4201–S4211. 10.21037/jtd.2018.08.56 [doi] PMID: 30687536

31. Sulit L, Storfer-Isser A, Kirchner HL, Redline S. Differences in polysomnography predictors for hypertension and impaired glucose tolerance. Sleep. 2006;29(6):777–783. 10.1093/sleep/29.6.777 [doi] PMID: 16796216

32. Kapur VK, Resnick HE, Gottlieb DJ, for the Sleep Heart Health Study Group. Sleep Disordered Breathing and Hypertension: Does Self-Reported Sleepiness Modify the Association? Sleep. 2008;31(8):1127–1132. 10.5665/sleep/31.8.1127

33. Mazzotti DR, Keenan BT, Lim DC, Gottlieb DJ, Kim J, Pack AI. Symptom Subtypes of Obstructive Sleep Apnea Predict Incidence of Cardiovascular Outcomes. Am J Respir Crit Care Med. 2019;200(4):493–506. 10.1164/rccm.201808-1509OC [doi] PMID: 30764637

34. Ogilvie RP, Lakshminarayan K, Iber C, Patel SR, Lutsey PL. Joint effects of OSA and self-reported sleepiness on incident CHD and stroke. Sleep Med. 2018;4432–37. S1389-9457(18)30016-9 [pii] PMID: 29530366

